# Patients’ satisfaction and quality of clinical laboratory services provision at public health facilities in northeast Ethiopia

**DOI:** 10.1101/2022.01.25.22269238

**Authors:** Daniel Dagne Abebe, Minwuyelet Maru Temesgen, Addisu Tesfie Abozin, Abebe Eyasu Zeleke, Seid Legesse Hassen, Hailay Berhe Gebremichael

**Affiliations:** Amhara Public Health Institute Dessie Branch, Dessie, Amhara region, Ethiopia; Amhara Public Health Institute Head Office, Bahir-dar, Amhara region, Ethiopia; ICAP Ethiopia, Regional Laboratory Advisor, Mekele, Tigray region, Ethiopia

**Keywords:** Medical Laboratories, Quality Care, Customer Satisfaction, Customer Perception, Patient Care Experience, Organizational Factors, Objective Quality Measures, Ethiopia

## Abstract

Patient satisfaction is a key element of quality measures that has increasingly become acknowledged as an important tool for service improvement. This study aimed to assess the level of patients’ satisfaction and associated factors with clinical laboratory services provided at public health facilities. A cross-sectional study was conducted from May-June 2019 among clients attending 24 health centers and 8 hospitals, northeast Ethiopia. A total of 502 patients were selected using systematic random sampling. Patient’s satisfaction towards multiple aspects of laboratory services was assessed using structured exit interview questionnaire, on a rating scale of 1 (very dissatisfied) to 5 points (very satisfied). We assessed test availability and laboratory practices using facility inventory, stepwise accreditation audit checklist and blinded slide rechecking. Data were entered and analyzed using EpiData ver3.1 and STATA ver14.1. Multivariable logistic regression analysis was used to determine the association of factors with overall satisfaction. Overall, majority of the respondents (73.5%) were found to be satisfied. Lowest mean ratings were obtained for waiting area (3.3), and information provided on specimen collection (3.5) and on how and when to receive results (3.7). Patients were more likely to be satisfied in health centers (75.2%) than in hospitals (68.6%) (AOR=1.9, 95%CI: 1.0-3.6, *p*=0.036). Patients’ timely receipt of results (*p*=0.005) and laboratories’ accuracy of results (*p*< 0.025) also showed significant positive associations with satisfaction. In conclusion, there were specific areas of deficiency that were driving dissatisfaction, particularly in the larger hospital laboratories. Therefore, more and balanced emphasis should be given to the patients’ experiences, alongside technical quality improvements, to reduce the disparities and enhance the overall quality of care.

## Introduction

Medical laboratory services are an essential component of strong national health systems. Laboratory diagnosis plays a crucial role for effective clinical management and control of major diseases of public health importance [1]. This requires quality diagnostic services to be available and properly utilized at different levels, and the test results to be accurate, reliable and timely [2,3]. However, despite the rapidly expanded access to health care, the quality of laboratory services has remained sub-optimal in many resource-limited countries like Ethiopia [4,5]. Where, laboratories are often faced with several challenges like poor infrastructure, lack of quality and competency among health professionals, and lack of standardized operations, hampering the quality care delivered to patients [6].

High-quality health care organizations strive to practice according to evidence-based standards, and thus meet the needs and expectations of patients, the ultimate beneficiaries of health services [7,8]. Patients’ satisfaction is a key aspect of health care quality that is being increasingly recognized for its importance [3,9]. Satisfied patients are more likely to return to the same facility; and this has direct impacts on customers’ loyalty, health insurance coverage and organizational sustainability. In today’s competitive environment, patient-centred care has got increasing focus as even the most technically competent care is meaningless if it does not satisfy the patients [2,3]. In addition, positive perceptions of care might often be translated into more positive adherence to recommended care and treatment; thereby enhancing patient health outcomes [10]. Quality management and accreditation standards also emphasize the importance of customers’ feedbacks in identifying opportunities for quality improvement [1,9].

In Ethiopia, laboratories are integrated structurally with the national health care system, which includes health centre, and primary, general and specialized hospitals [4,5]. Parallel to the rapidly expanded access to health care in the country, there is a growing need to improve quality of services [4,5,11]. However, availability of quality laboratory diagnostic services has been reported to remain suboptimal [12,13], where more than half of the laboratories were not capable of conducting all basic tests expected. Considerable efforts, including the WHO/AFRO’s stepwise accreditation, have been undertaken to strengthen quality systems of laboratories in such resource-limited countries of Africa [1,4]. However, the actual impact on quality of care and thus patient satisfaction has been less explored in local settings [6,14-16]. Laboratories are required to conduct customer satisfaction assessments to maintain their accreditation status in developed countries, yet not common in developing countries.

Quality health care is commonly viewed as having three closely inter-related dimensions: structural, process (inter-personal and technical), and outcome quality [8,17,18]. Where, patient satisfaction is acknowledged as an indispensable element of outcome quality measures, besides clinical health outcomes [7,19]. Insight into the different components is essential to efficiently detect quality problems, and this needs evaluating quality from multiple perspectives [7,20]. Two distinct approaches have been often employed in assessing quality performance: direct assessments and customer satisfaction surveys. A set of objective key performance indicators are used to directly measure specific areas of clinical practice as compared to published standards [1,21]. Customers’ satisfaction is a measure of the perceived quality of care received by patients as compared to the care expected by patients [20]. Different quality attributes have been considered as relevant determinants for measuring overall satisfaction, such as access to care, physical environment, readiness of services, quality system and reliability of the clinical or diagnostic care [8,20,22]. However, clients might fail to effectively discriminate between services that are different in quality, especially in technical quality, as they might often be more sensitive to inter-personal or tangible aspects [20,23-26].

A number of factors have been reported to influence the patients’ satisfaction with laboratory services, including socio-demographic characteristics like age, education and severity of illness [27,28], and patients’ visit-related experiences like turnaround time of results (TAT) [22,29], phlebotomy skills [13] and facility settings [30]. However, available evidence has been unclear on the correlation of patients’ experiences with objective measures of quality practice, and thus the reliability of patient-based measures remain debated [10,24,25,31]. Particularly, the previous studies conducted in Ethiopia and other resource-limited settings have rarely examined specific organizational factors that may drive satisfaction of laboratory users [14,15,22,30]. Comprehensive understanding of determinants at individual- and organizational-levels would provide more opportunities for improving service delivery and satisfaction in local settings [30,32]. Therefore, the aim of this study was to assess patients’ satisfaction level and associated patient- and organizational-level factors at clinical laboratories of public health facilities in a resource-limited setting.

## Materials and methods

### Study design and area

A facility-based cross-sectional study was conducted from May to June, 2019 in east Amhara region, northeast Ethiopia. This area covers six zonal administrations, comprising about half of the Amhara region. There were 35 public hospitals and 367 health centers (402 health facilities total) at the time of the study. Out of these, 252 (35 hospitals and 217 health centers) had functional laboratory diagnostic service. The health facilities provide different clinical and laboratory services to the community, including basic tests like stool examination, urinalysis, serology, gram stain, malaria and tuberculosis (TB) microscopy, and manual haematology and chemistry tests [33]. Additional services provided at hospital levels include more advanced tests like automated clinical chemistry and haematology tests, CD4 count, electrolyte, hormone analysis, and microbiology tests [33]. Amhara Public Health Institute Dessie branch (APHIDB), located at Dessie, is responsible to coordinate capacity building and external quality assurance (EQA) activities to all laboratories in east Amhara, in collaboration with partners.

### Source population

All adult patients (aged ≥18 years) who were using clinical laboratory services at public health facilities of east Amhara, northeast Ethiopia were source population.

### Study population and eligibility criteria

Adult patients who received general laboratory services at the randomly selected government health facilities during the study period were study population. Those patients who were critically ill and unable to respond and those not voluntary to participate were excluded.

### Sample size and sampling procedure

A total of 32 health facilities (24 health centers and 8 hospitals) were selected and included, accounting for 13% of the total 252 diagnostic public health facilities found in the study area. The sample size of clients was determined using the common formula for a single population proportion, n = (Z^2^_1-α/2_) *p* (1-p)/d^2^. Considering a 95% confidence level (Z_1-α/2_ = 1.96), p= 78.6% satisfaction level from a study done in Ethiopia nationally [22], a margin of error of d= 5%, and then multiplying by a design effect of 2.0 as stratified sampling of facilities. Therefore, considering a 5% non-response rate, the final sample size of patients required was determined to be **<u>n= 536 patients</u>**. The total sample size of clients was then pre-allocated to each facility proportional to facility type’s average client flow for operational feasibility-15 clients from a health center and 22 from a hospital.

Facilities were first selected using stratified systematic random sampling method with probability proportionate to facility’s size (estimated monthly client volume). Updated list sampling frame of facilities was prepared in geographical order providing implicit stratification (by facility type and zone), using Excel spreadsheet [34]. Systematic random sampling was used to select the 32 facilities at once, using a single sampling interval (k) determined based on cumulative facility sizes. At each facility, patients were then selected using systematic random sampling from clients consecutively visiting the laboratory during the study period (two days per facility). Sampling intervals (k) of clients varied based on each facility’s expected client flow. This ensures sufficient sample from stratum with fewer number of facilities but larger client sizes, while maintaining the final client sample self-weighting.

### Study variables

The dependent variable was the overall satisfaction level of patients, based on their perceptions towards multiple aspects of laboratory services (eg. cleanliness, adequacy of waiting area, information provision). Patient-level independent variables include socio-demographic (eg. age, sex, education, residence) and patient-reported experiences of care (eg. accessibility of service locations, complete availability of ordered tests, receipt of results within TAT, encounter of results lost, affordability). Organizational-level variables include facility type, and objective measures of laboratory practice related to structure (eg. test menu capacity) and technical process (eg. stepwise accreditation level, accuracy of results).

### Data collection tools and procedures

A pre-tested, structured and interviewer-administered questionnaire was used for collecting data from exiting patients. The questionnaire contained different types of questions related to socio-demographic and visit related characteristics (8), patient-reported experiences of care (14), and levels of satisfaction (12). Standardized five-point Likert rating scale based responses ranging from “very dissatisfied” to “very satisfied” (1 to 5 points) were used for each of the 12 satisfaction measuring items.

Facility assessments were conducted for collecting organizational-level data and evaluate laboratory practices based on selected objective key performance indicators. Structured laboratory inventory checklist was used to assess readiness and functionality of ranges of expected testing services, as per the regional laboratory standard [33]. Stepwise accreditation audit checklist of WHO/AFRO was adopted to evaluate laboratory practices and quality systems against evidence-based standards [1]. Blinded slide rechecking was also performed to assess reliability of diagnosis results, by systematically collecting a representative sample of malaria and TB microscopy slides examined in the last three months [35,36].

The patient exit interview questionnaire was customized from existing tool validated for customers of general laboratory nationally [22]. The English version of the questionnaire was translated into local language (Amharic) for use in local settings. Data collectors were senior laboratory experts recruited from external facilities. Data collectors and supervisors were then trained on the data collection tools and procedures. All tools were also pre-tested in nearby facilities not included in the study and customized as needed, before use for the actual data collection.

### Measurements

Satisfaction ratings given for the twelve satisfaction measuring questions were averaged to create an overall mean satisfaction (weighted average) score for each respondent. Patients with satisfaction score of ≥4 out of five points were classified as satisfied (i.e., very satisfied plus satisfied ratings), while the rest were classified as not satisfied. For each laboratory, adequacy of test menu capacity was measured as the percentage of test menu available and provided on the day of visit out of the standard test menu expected for the respective facility type. The standard recommends 37 tests for a health centre, and 63, 90 or 120 tests for primary, general or specialized hospitals, respectively [33]. Service interruption period was measured as percentage of non-functionality period due to stock-out or equipment failure within the previous three months [33]. Stepwise accreditation score was calculated as percentage of points met out of the total points of the audit checklist (275 points); and star grades were assigned for each laboratory starting from zero star (if score <55%) up to five star (if score ≥95%) [1]. Concordance rates for malaria and TB diagnosis results were calculated as percentage of correct readings out of the total slides rechecked for each facility. For this, 30 malaria and 40 TB microscopy slides were collected per facility in average, following recommendations of EQA guidelines [35,36].

### Data quality assurance

Training was given for data collectors and supervisors on recruitment of participants and data collection. Pre-tested, structured and standardized tools were used to ensure consistency of data collection. Certified external laboratory assessors conducted the laboratory assessments. Supervisors and research team supervised the data collection and checked consistency and completeness of filled questionnaires. Interviews were conducted in a location where the facility staff and other patients were not present.

### Operational definition

***Satisfaction level:*** is patient’s perception or feeling of the degree to which service quality attributes have fulfilled customer’s needs and requirements, and is acknowledged as a measure of outcome quality in this study.

### Data management and analysis

Data was entered using EpiData version 3.1 (for patient data) and MS Excel 2010 (for facility data), and all data was exported to STATA version 14.1 for main analysis. Descriptive statistics (mean, SD, and proportions) were computed and compared by facility type. We used stepwise multivariable logistic regression analysis to identify individual- and organizational-level factors affecting the patients’ overall satisfaction level (as categorical variable). Robust standard errors were applied in order to account for the clustering of patients within facilities. Those variables with a *p*-value <0.20 in initial bivariate analysis were considered to be included in to the multivariable model. A *p*-value <0.05 was considered statistically significant. Adjusted odds ratios (AORs) with 95% confidence intervals (CIs) were reported.

### Ethical consideration

Ethical clearance was obtained from the regional Ethical Review Board of Amhara Public Health Institute (APHI) Head office, located at Bahir-dar, Amhara region, Ethiopia. The respective zonal health departments and facility administrators were informed about the general aim and significance of the study through an official permission letter. The general aim and purpose of the study was described to each eligible patient and all voluntary participants gave verbal informed consent prior to enrolment. Participants were also informed about their right to refuse at any time and no consequences or risks on them due to expressing any dissatisfaction. Data was collected anonymously, without personal identifiers to ensure confidentiality.

## Results

### Patients’ socio-demographic characteristics

A total of 502 patients participated in this study with a response rate of 93.7%. Table 1 shows the socio-demographic characteristics of the patients. The mean age (±SD) of participants was 35.4 (±13.7) years, and more than third of them (36.9%) were farmers. The mean distance (±SD) travelled by patients to reach the facility was 18.4Km (±46.7), ranging from less than 1Km to 500Km. When compare the distribution of patients by facility type, the mean distance (±SD) travelled by patients and the percentage of first-time visitors were found to be significantly higher for hospitals (39.2 ±67.8, 60.3%) than that of health centres (9.8Km ±30.8, 38.3%) (both, *p*= 0.000), respectively.

**Table 1.**
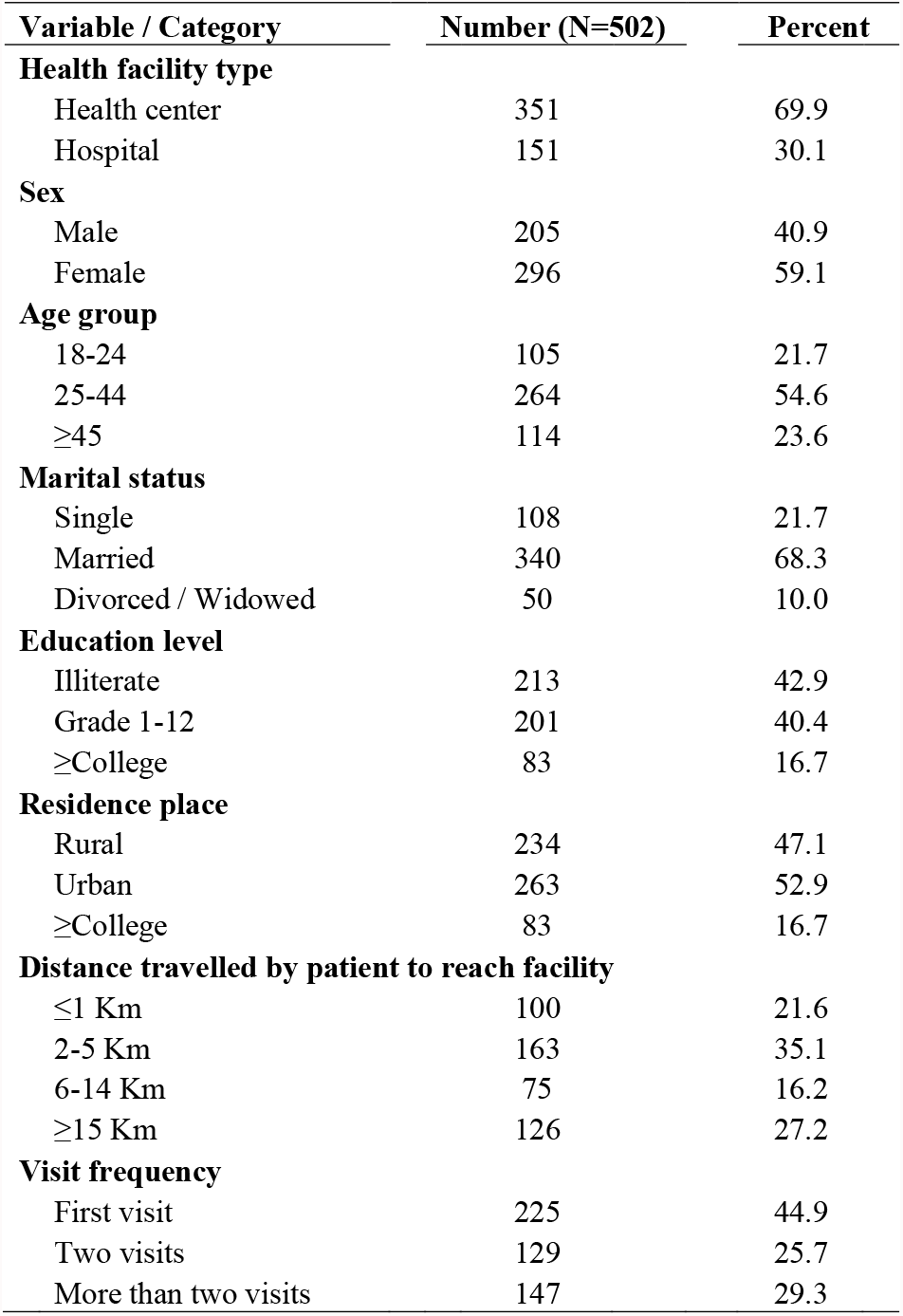
Patients’ socio-demographic characteristics at selected health facilities, northeast Ethiopia, 2019.

### Patient-reported experiences of laboratory services

Majority (77.2%) of the patients reported not to get a place to put personal things (like coat, bag, etc.) during blood drawing (Table 2). More than one quarter (27.2%) of the patients reported not to receive results within the claimed TAT, and 15.3% encountered result lost. As compared to health centers, hospitals had more percentages of patients who did not receive results within the claimed TAT, encounter results lost, personnel did not ensure bleeding stopped before releasing patient (all, *p*= 0.000), or encountered difficulty of getting locations of laboratory (*p*= 0.000) or cashier rooms (*p*= 0.032).

**Table 2.** Patients’ experiences of laboratory services by facility type, northeast Ethiopia, 2019 (N= 351 patients total, 351 from health centers, 151 from hospitals) ^a^.

|  | Patient Yes n (%) |  |  |  |  |  |  |  |
| --- | --- | --- | --- | --- | --- | --- | --- | --- |
| Patients reported (Yes) | Health center |  | Hospital |  | Total |  | Chi<sup>2</sup> | p |
| Got location of laboratory easily | 337 | 96.6 | 123 | 81.5 | 460 | 92.0 | 32.7 | 0.000 |
| Got location of cashier easily | 290 | 90.9 | 122 | 84.1 | 412 | 88.8 | 4.6 | 0.032 |
| Got location of latrine easily | 231 | 88.5 | 102 | 85.7 | 333 | 87.6 | 0.6 | 0.443 |
| Personnel put on fresh gloves | 281 | 87.8 | 127 | 87.6 | 408 | 87.7 | 0.0 | 0.945 |
| Personnel labeled samples | 239 | 73.1 | 122 | 85.3 | 361 | 76.8 | 8.4 | 0.004 |
| Place available to put personal things at phlebotomy area<sup>a</sup> | 49 | 23.8 | 20 | 20.6 | 69 | 22.8 | 0.4 | 0.540 |
| Needle stick attempts during blood drawing ≥two times<sup>a</sup> | 21 | 9.6 | 13 | 12.1 | 34 | 10.4 | 0.5 | 0.478 |
| Develop bruise after phlebotomy | 44 | 20.8 | 23 | 21.5 | 67 | 21.0 | 0.0 | 0.878 |
| Bleeding stopped before release<sup>a</sup> | 200 | 95.2 | 82 | 77.4 | 282 | 89.2 | 23.5 | 0.000 |
| Result received as claimed TAT | 273 | 78.9 | 69 | 55.6 | 342 | 72.8 | 24.9 | 0.000 |
| Encountered results lost/missing | 30 | 8.8 | 45 | 30.0 | 75 | 15.3 | 36.2 | 0.000 |
| Got complete tests ordered | 300 | 92.3 | 140 | 94.6 | 440 | 93.0 | 0.8 | 0.365 |
| Service charge / price fair | 242 | 79.6 | 121 | 92.4 | 363 | 83.4 | 10.8 | 0.001 |

<sup>a</sup>Total respondent not equal to total sample size for these variables due to non-applicable question and/or missing responses.

### Characteristics of participated health facility laboratories

Twenty four health centres (75.0%) and eight hospitals (25.0%) were included in this study. Number of test menu capacity increased with increase from health centers to larger hospitals (Table 3 and Fig 1). However, only 15.8% of the health centers and 37.5% of hospitals provided ≥75% of the expected test menu as per the standard. Majority (87.5%) of the laboratories achieved zero star (i.e., score <55%) in implementing stepwise accreditation, while the remainder (12.5%) achieved one or two star grades. More percentage of hospitals achieved one or two star grades (37.5%) as compared to that of health centres (4.2%) (*p*= 0.024). Among the twelve quality system components, the lowest mean scores were observed for identification of non-conformities, evaluation, and continuous improvement (range from 6.2%±9.1 to 21.4%±26.1). Finally, the rate of agreement of malaria species diagnosis results showed huge variation across individual laboratories (range 25.0%-100.0%), and showed a tendency to decrease for hospitals (84.3% ±25.7) than that of health centres (94.8% ±10.1) (*p*= 0.157).

**Table 3.** Descriptive summary of laboratory performance indicators by facility type, northeast Ethiopia, 2019 (N= 24 health centers, 8 hospitals).

| Indicator | Mean score (± SD) |  |  |  |  |  | *p*<sup>a</sup> |
| --- | --- | --- | --- | --- | --- | --- | --- |
|  | Health center |  | Hospital |  | Overall |  |  |
| Total number of test menu provided | 20.7 | 5.6 | 50.8 | 15.9 | 57.3 | 12.9 | 0.000 |
| Percentage of test menu provided | 53.0 | 14.4 | 56.4 | 20.9 | 54.0 | 16.2 | 0.631 |
| Service uninterrupted period | 99.0 | 2.2 | 99.5 | 0.5 | 99.1 | 1.9 | 0.547 |
| Standard service availability period | 52.5 | 14.5 | 56.1 | 20.8 | 53.6 | 16.3 | 0.612 |
| Stepwise accreditation audit score | 27.4 | 12.9 | 49.9 | 13.6 | 33.0 | 16.2 | 0.000 |
| Concordance rate of malaria microscopy | 98.6 | 3.3 | 97.2 | 3.1 | 98.2 | 3.3 | 0.331 |
| Species agreement rate of malaria | 94.8 | 10.1 | 84.2 | 25.7 | 91.3 | 17.1 | 0.157 |
| Concordance rate of TB microscopy | 98.0 | 5.2 | 99.6 | 0.7 | 98.3 | 4.7 | 0.533 |

<sup>a</sup>*p*-value from *t*-test comparing mean scores by facility type (health center, hospital).

**Figure 1.**
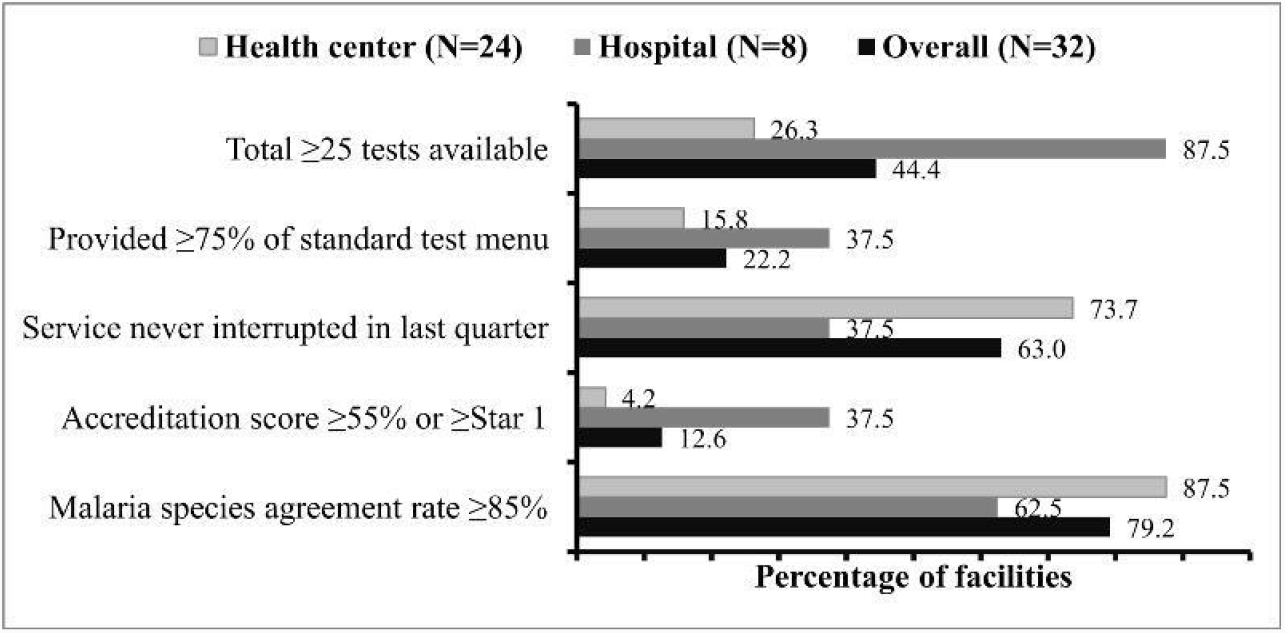
Proportions of facilities with observed high levels of laboratory performance indicators by facility type.

### Patients’ satisfaction level with clinical laboratory services

The overall percentage of patients satisfied with the clinical laboratory services received was 73.5% (95%CI: 71.8-75.7) (Table 4). In Likert rating scale, the overall mean score of satisfaction was 3.81 ±0.49, with ratings for specific aspects ranging from 3.3 to 4.2. Lowest mean ratings of satisfaction were obtained for the cleanness of latrines (3.3), adequacy of waiting area (3.4), and provision of information on when and how to receive results (3.5) and on procedures of specimen collection (3.7). As compared to health centres (75.2%), hospitals had a lower percentage of overall satisfied patients (68.6%) (*p*= 0.002) (Fig 2). Specific areas of significant difference with more dissatisfaction for the larger hospitals included all of the lowest rated aspects mentioned (all, *p*< 0.016).

**Table 4.** Patient satisfaction levels towards different aspects of laboratory services, northeast Ethiopia, 2019 (N= 502 patients)^a^.

|  | Number (%) |  |  |  |  |  |  |  |  |  | Mean | SD | %Satisfied<sup>b</sup> |  |
| --- | --- | --- | --- | --- | --- | --- | --- | --- | --- | --- | --- | --- | --- | --- |
| Satisfaction items | Very dissatisfied |  | Dissatisfied |  | Neutral |  | Satisfied |  | Very satisfied |  |  |  | n | % |
| Cleanliness of waiting area | 22 | 4.4 | 24 | 4.8 | 86 | 17.2 | 290 | 58.0 | 78 | 15.6 | 3.8 | 0.9 | 368 | 73.6 |
| Adequacy of waiting area | 60 | 12.0 | 64 | 12.8 | 66 | 13.2 | 256 | 51.2 | 54 | 10.8 | 3.4 | 1.2 | 310 | 62.0 |
| Availability of staff on work | 12 | 2.4 | 28 | 5.6 | 58 | 11.6 | 299 | 59.8 | 103 | 20.6 | 3.9 | 0.9 | 402 | 80.4 |
| Information at reception | 14 | 2.8 | 27 | 5.4 | 77 | 15.4 | 297 | 59.3 | 86 | 17.2 | 3.8 | 0.9 | 383 | 76.4 |
| Appearance of employees | 1 | 0.2 | 4 | 0.8 | 50 | 10.0 | 292 | 58.2 | 155 | 30.9 | 4.2 | 0.7 | 447 | 89.0 |
| Courtesy/respect of staff | 11 | 2.2 | 13 | 2.6 | 85 | 17.0 | 267 | 53.4 | 124 | 24.8 | 4.0 | 0.8 | 391 | 78.2 |
| Privacy assurance measure<sup>a</sup> | 4 | 0.9 | 15 | 3.4 | 57 | 13.0 | 225 | 51.3 | 138 | 31.4 | 4.1 | 0.8 | 363 | 82.7 |
| Cleanness of specimen area<sup>a</sup> | 5 | 1.1 | 10 | 2.1 | 78 | 16.6 | 262 | 55.9 | 114 | 24.3 | 4.0 | 0.8 | 376 | 80.2 |
| Information provided on specimen collection<sup>a</sup> | 12 | 3.1 | 23 | 5.9 | 83 | 21.2 | 218 | 55.8 | 55 | 14.1 | 3.7 | 0.9 | 273 | 69.8 |
| Cleanliness of latrine<sup>a</sup> | 49 | 12.9 | 50 | 13.2 | 87 | 23.0 | 140 | 36.9 | 53 | 14.0 | 3.3 | 1.2 | 193 | 50.9 |
| Information on when/how to receive results | 25 | 5.1 | 48 | 9.8 | 119 | 24.4 | 226 | 46.3 | 70 | 14.3 | 3.5 | 1.0 | 296 | 60.7 |
| General quality of service | 3 | 0.6 | 11 | 2.3 | 106 | 22.1 | 286 | 59.6 | 74 | 15.4 | 3.9 | 0.7 | 360 | 75.0 |
| Overall satisfaction <sup>c</sup> | - | - | - | - | - | - | - | - | - | - | 3.8 | 0.5 | 369 | 73.5 |

<sup>a</sup> Total may not add up to total sample size for these variables due to question non-applicable and/or missing response.
<sup>b</sup> Percentage of satisfied patients calculated defining very satisfied plus satisfied ratings as satisfied. (i.e., using a Likert scale score of $\geq 4$ ).
<sup>c</sup> Overall satisfaction (weighted average) score was measured as average based on the twelve satisfaction items.

**Figure 2.**
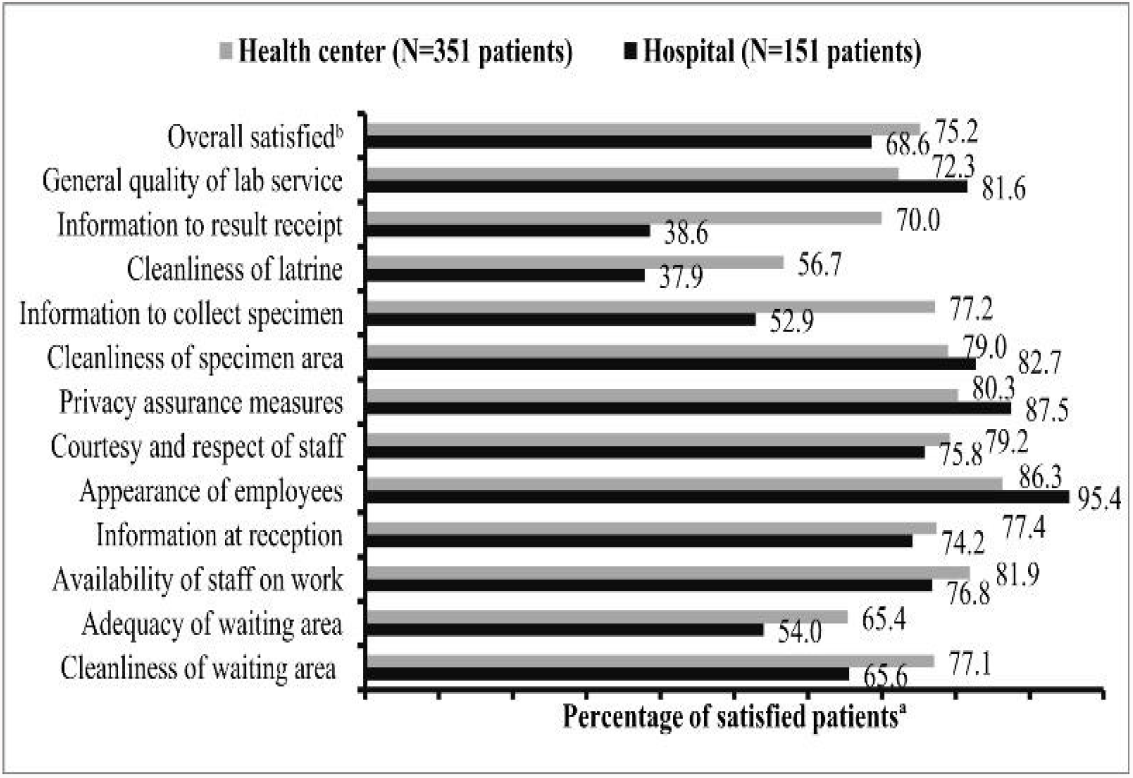
Percentages of satisfied patients for different aspects of clinical laboratory services by facility type. ^a^Satisfied defined as very satisfied plus satisfied responses, i.e., at a score of ≥4 as cut-off. ^b^Overall satisfied patients defined at ≥4 of the “weighted average score” of the twelve items.

### Factors associated with patient satisfaction

On multivariable analysis, the patients’ distance travelled, receipt of results within the claimed TAT and personnel put on fresh gloves, as well as the laboratories’ facility type and concordance rates of TB and malaria species diagnosis results were found to have significant association with the overall level of patients’ satisfaction (all, *p*< 0.05) (Table 5). Hospital patients were about two times more likely to be dissatisfied with laboratory services than those of health centers (AOR= 1.9; 95%CI: 1.0-3.6; *p*= 0.036). Patients who reported personnel to wear fresh gloves were also about three times more likely to be satisfied than those who reported personnel not to wear fresh gloves (AOR= 2.5; 95%CI: 1.2-5.2; *p*= 0.012).

**Table 5.**
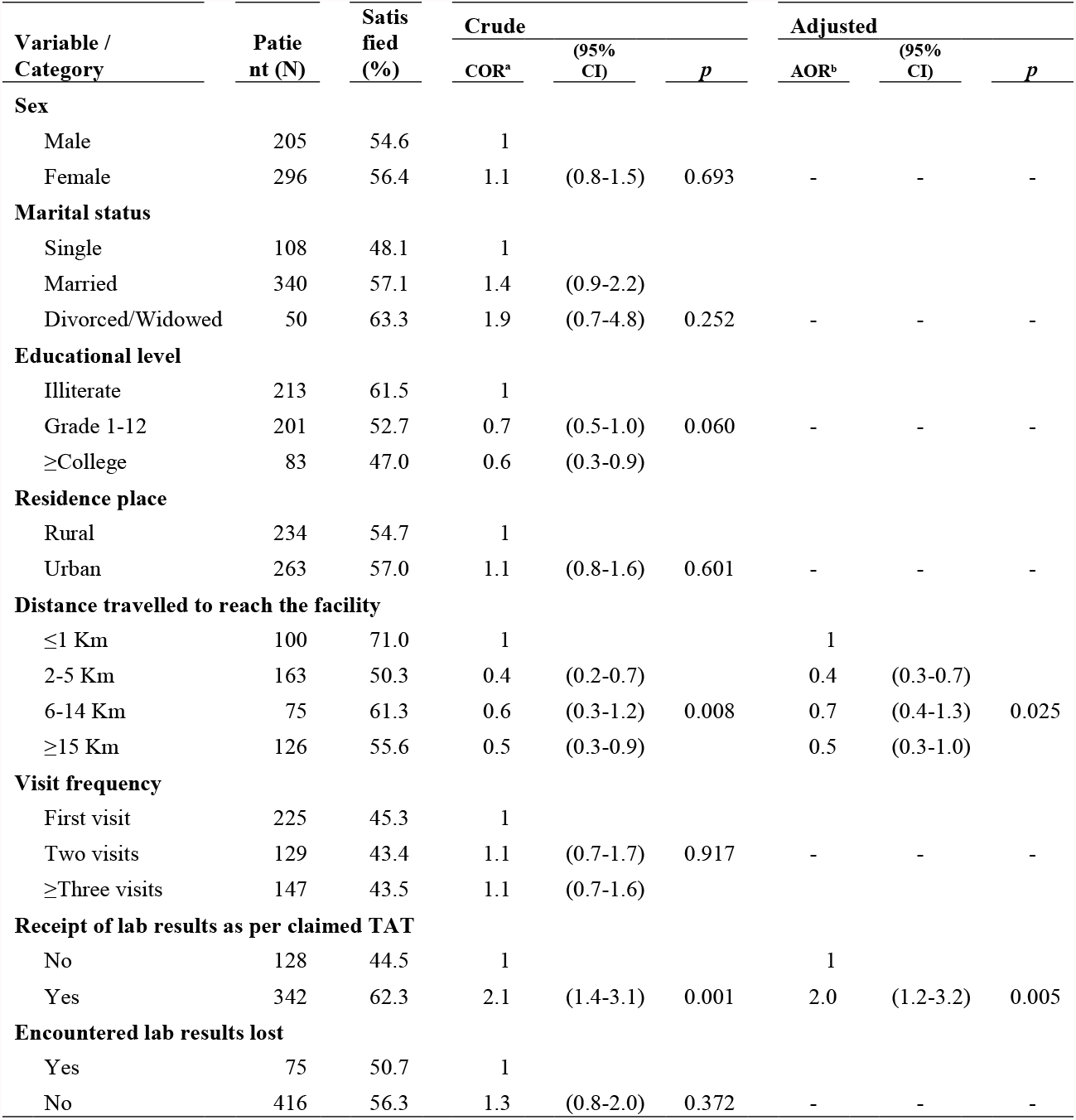
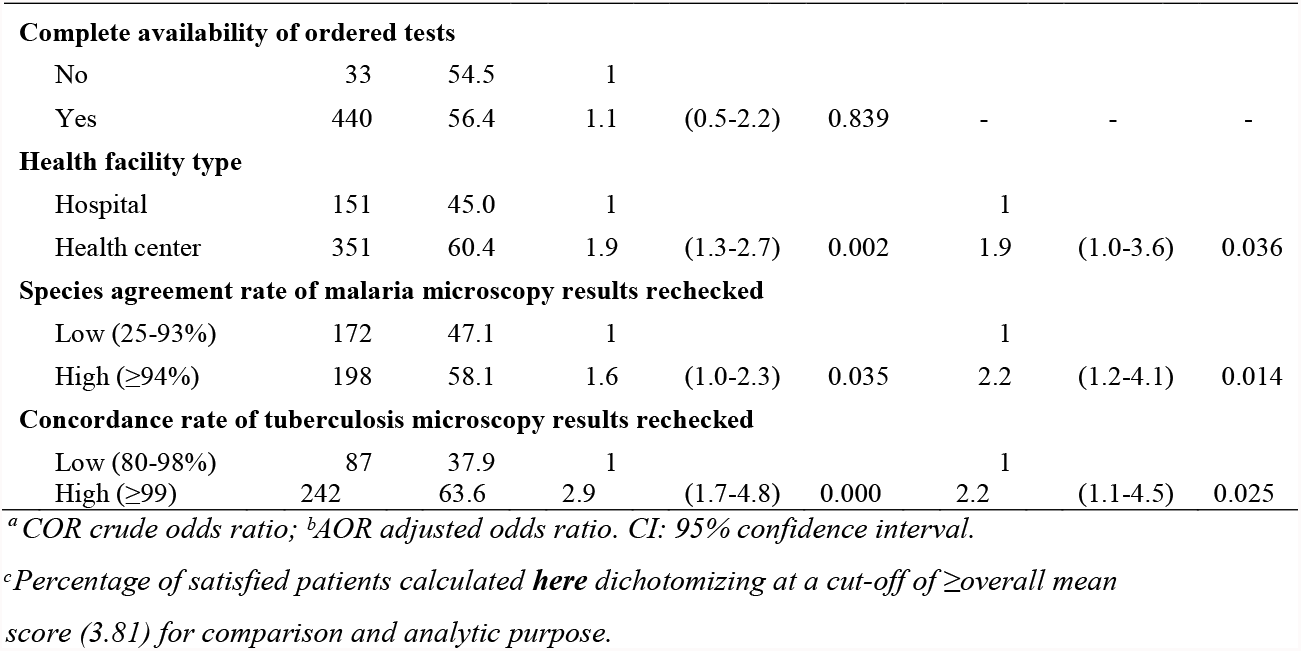
Association of independent variables with satisfaction level of patients at public health facilities in northeast Ethiopia, 2019.

| Variable /<br>Category | Patie<br>nt (N) | Satis<br>fied<br>(%) | Crude |  |  | Adjusted |  |  |
| --- | --- | --- | --- | --- | --- | --- | --- | --- |
|  |  |  | COR<sup>a</sup> | (95%<br>CI) | *p* | AOR<sup>b</sup> | (95%<br>CI) | *p* |
| Sex |  |  |  |  |  |  |  |  |
| Male | 205 | 54.6 | 1 |  |  |  |  |  |
| Female | 296 | 56.4 | 1.1 | (0.8-1.5) | 0.693 | - | - | - |
| Marital status |  |  |  |  |  |  |  |  |
| Single | 108 | 48.1 | 1 |  |  |  |  |  |
| Married | 340 | 57.1 | 1.4 | (0.9-2.2) |  |  |  |  |
| Divorced/Widowed | 50 | 63.3 | 1.9 | (0.7-4.8) | 0.252 | - | - | - |
| Educational level |  |  |  |  |  |  |  |  |
| Illiterate | 213 | 61.5 | 1 |  |  |  |  |  |
| Grade 1-12 | 201 | 52.7 | 0.7 | (0.5-1.0) | 0.060 | - | - | - |
| ≥College | 83 | 47.0 | 0.6 | (0.3-0.9) |  |  |  |  |
| Residence place |  |  |  |  |  |  |  |  |
| Rural | 234 | 54.7 | 1 |  |  |  |  |  |
| Urban | 263 | 57.0 | 1.1 | (0.8-1.6) | 0.601 | - | - | - |
| Distance travelled to reach the facility |  |  |  |  |  |  |  |  |
| ≤1 Km | 100 | 71.0 | 1 |  |  | 1 |  |  |
| 2-5 Km | 163 | 50.3 | 0.4 | (0.2-0.7) |  | 0.4 | (0.3-0.7) |  |
| 6-14 Km | 75 | 61.3 | 0.6 | (0.3-1.2) | 0.008 | 0.7 | (0.4-1.3) | 0.025 |
| ≥15 Km | 126 | 55.6 | 0.5 | (0.3-0.9) |  | 0.5 | (0.3-1.0) |  |
| Visit frequency |  |  |  |  |  |  |  |  |
| First visit | 225 | 45.3 | 1 |  |  |  |  |  |
| Two visits | 129 | 43.4 | 1.1 | (0.7-1.7) | 0.917 | - | - | - |
| ≥Three visits | 147 | 43.5 | 1.1 | (0.7-1.6) |  |  |  |  |
| Receipt of lab results as per claimed TAT |  |  |  |  |  |  |  |  |
| No | 128 | 44.5 | 1 |  |  | 1 |  |  |
| Yes | 342 | 62.3 | 2.1 | (1.4-3.1) | 0.001 | 2.0 | (1.2-3.2) | 0.005 |
| Encountered lab results lost |  |  |  |  |  |  |  |  |
| Yes | 75 | 50.7 | 1 |  |  |  |  |  |
| No | 416 | 56.3 | 1.3 | (0.8-2.0) | 0.372 | - | - | - |
| **Conti...Table 5:** |  |  |  |  |  |  |  |  |
| **Complete availability of ordered tests** |  |  |  |  |  |  |  |  |
| No | 33 | 54.5 | 1 |  |  |  |  |  |
| Yes | 440 | 56.4 | 1.1 | (0.5-2.2) | 0.839 | - | - | - |
| **Health facility type** |  |  |  |  |  |  |  |  |
| Hospital | 151 | 45.0 | 1 |  |  | 1 |  |  |
| Health center | 351 | 60.4 | 1.9 | (1.3-2.7) | 0.002 | 1.9 | (1.0-3.6) | 0.036 |
| **Species agreement rate of malaria microscopy results rechecked** |  |  |  |  |  |  |  |  |
| Low (25-93%) | 172 | 47.1 | 1 |  |  | 1 |  |  |
| High (≥94%) | 198 | 58.1 | 1.6 | (1.0-2.3) | 0.035 | 2.2 | (1.2-4.1) | 0.014 |
| **Concordance rate of tuberculosis microscopy results rechecked** |  |  |  |  |  |  |  |  |
| Low (80-98%) | 87 | 37.9 | 1 |  |  | 1 |  |  |
| High (≥99) | 242 | 63.6 | 2.9 | (1.7-4.8) | 0.000 | 2.2 | (1.1-4.5) | 0.025 |

<sup>a</sup>COR crude odds ratio; <sup>b</sup>AOR adjusted odds ratio. CI: 95% confidence interval.
<sup>c</sup>Percentage of satisfied patients calculated **here** dichotomizing at a cut-off of ≥overall mean score (3.81) for comparison and analytic purpose.

## Discussion

This study assessed patients’ perceptions towards clinical laboratory services’ quality, and examined the effect of both individual- and organizational-level factors in a resource-limited setting, northeast Ethiopia. It has been revealed that specific gaps related to both patient experience and technical process quality play a role in driving dissatisfaction, especially in the larger hospitals. However, the patients’ socio-demographic characteristics (except distance) and inter-personal quality aspects, and the laboratories’ structural diagnostic capacity and stepwise accreditation levels were revealed to have a lesser or no role.

Overall, the majority (73.5%) of patients appeared to be satisfied with the laboratory services received (Table 4), which was consistent with finding from Ethiopia (73.0%-77.1%) [29,37]. The finding was higher than previous finding of most studies from the same country (54.1% to 60.4%) [13,28,38-41]. However, cautions should be considered when interpreting such client-based findings in general, as client feedbacks often tend to be positive due to social desirability bias or lack of technical expertise. On the other hand, the finding appeared lower when compared with finding of other studies from Ethiopia (78.6%-90.8%) [22,42-44]. However, the previous studies assessed specific laboratory services with more emphasis by vertical programs, employed a lower demarcation threshold, or excluded mid-point or neutral ratings (with a score of 3 point) from the denominator when dichotomizing satisfaction. Huge variations exist in the demarcation threshold techniques employed among existing literature, making percentage-based comparisons difficult. Probably, the more direct summary measures, like weighted average score, should be encouraged for such Likert scale-based data to be reported always.

Lowest satisfaction ratings were obtained for sanitation of latrines and sitting arrangements of waiting areas followed by the provision of instructions on specimen collection and on result receipt (Table 4). This reflect the poor attention often given by facility administrators and providers for physical environment [22,37,40,42] and information provision [22,39,44]. These problems were especially more aggravated in the larger hospitals (Fig 2), which might also be in part related to the higher number of clients or deficiency of time to provide explanations in such settings [28,40]. Failure to communicate relevant information to patients could result in poor quality of specimens to be submitted by patients, or could leave users with biased expectations of TATs leading to customer dissatisfaction. Therefore, providers should improve their attitudes on the role of providing clear and complete information and advisory services to customers, that would improve quality of the technical output as well as the users’ experience.

Receipt of results within the claimed TAT showed significant independent effect on the overall satisfaction level of patients (Table 5), in line with findings of many studies done in Ethiopia [11,22,29,38,40,42,45]. More percentage of patients reported to receive results out of the claimed TAT (45.4%) and to encounter results lost (30.0%) in the larger hospitals (Table 2). This could be due to inefficient systems and work processes especially in larger organizations with heavy workloads, or biased expectations of clients with lack of awareness on agreed TATs, as mentioned above. Delayed services could delay clinical decision making besides distressing customers, and thus impacting patient health outcomes besides service utilization. Therefore, targeted interventions should consider redesigning work flows and availing improved technologies for efficient handling and communication of laboratory information. Laboratory providers need also to establish acceptable TATs for all test menu, clearly post and communicate it to customers, and monitor routine performance for continuous improvement.

The patients’ satisfaction level showed significant reduction with increase across facility levels (Table 5), revealing that the larger hospitals are not providing quality laboratory services as perceived by patients. As compared to primary health centres, larger facilities with better structural diagnostic capacities (Table 3 and Fig 1) were rather more likely to deliver poor actual experiences (Table 2) and satisfaction ratings (Fig 2). The finding suggests the bare availability of more advanced facilities may be less likely to guarantee improved perceptions of care [22,46,47], which is in contrast with previous findings [13,24,48-50]. The lower satisfaction could be explained by the biased or higher expectation of clients from the higher level hospitals, or by the actual poor quality of care in specific aspects by the existing larger hospitals with higher client loads. The other explanation could be in part due to the focus of the current policies of the country to strengthen primary health care services that are more accessible and familiar to the communities [46]. Given those hospital patients are often referral cases with critical conditions that may require urgent or advanced care, the poor acceptability of the care may be playing a big role in forcing people to prefer costly private providers [47]. The poor quality of the care may also be playing a role in health outcomes, as such patients are often at risk of more detrimental consequences. Therefore, clients should be encouraged to utilize primary level facilities for their routine care, while ensuring availability of comprehensive health care services at such community levels. Probably, there may also be a need to redesign the standard ranges of services so that the higher level hospitals will focus solely on more advanced and emergency care services, that may facilitate efficiency and continuity of care delivery [46].

The patients’ satisfaction level showed significant positive relationship with technical accuracy of the laboratory diagnosis results, as measured by blinded slide rechecking (Table 5). The observed accuracy of laboratory results tended to increase for the primary health centres (Table 3 and Fig 1), where better patient experiences (Table 2) and satisfaction ratings were obtained (Table 4), as mentioned. Evidence from existing literature remain argued on the reliability of clients’ judgements to reflect the actual quality of care, as clients might be more sensitive to behavioural or tangible aspects rather than clinical quality [23-25]. However, for example, providers with good technical competence could be more likely to be confident and thus to have good inter-personal qualities [26]. The current findings support evidence that suggest patient experiences to be closely linked with objective technical measures [10,31,51] validating customer feedbacks as meaningful and appropriate complements for organizational quality improvement. On the other hand, the observed mean rate of agreement in malaria species identification was unacceptable particularly for larger hospitals (84.2% ±25.7); which was lower when compared with previous reports from the same region (94.6%-96.6%) [35,36]. This problem could be related to poor technical competence of providers, or deficiency of time and negligence to examine sufficient fields due to work overloads.

We did not find association between the laboratories’ stepwise accreditation or star grade levels and the overall level of patients’ satisfaction (Table 5). Better quality systems were observed in the larger hospital laboratories (Table 3 and Fig 1), where patient experiences (Table 2) and satisfaction ratings were rather poorer (Table 4), as mentioned above. In contrast to previous findings [15,52], this finding supports questions raised on the claimed benefits of stepwise accreditation to improve quality of care and thus patient satisfaction especially in resource-limited settings [14,16,48]. Accreditation might be critiqued for the more focus on structure and process rather than customer focus and patient outcomes, and the demand for huge paper work [14,16,48], which may need further investigation. The lack of impact visible to patients in the present study setting could be explained in part by the fact that the overall implementation level was in infancy stage (Table 3 and Fig 1); or the lowest scores were observed particularly for measurement and continuous improvement related activities, which may be the key drivers for change.

Several limitations of this study should be kept in mind. First, the patients were recruited from the health facilities, not the communities. Such clients might tend to conceal much of their dissatisfactions due to social desirability bias. Second, a sample 12 size of 32 government facilities was not big enough to further compare between different hospital levels, and a survey of patient customers may not fully reflect the views of other stakeholder groups. Third, there might be other important variables that we did not consider in the present study, such as the patient’s severity of illness and workloads of the facilities. Further larger studies are thus needed to expand the scope to include private laboratories and other customer groups like physicians and the laboratory providers, themselves. Finally, client-based evaluations like most human judgments may not also be fully reliable to reflect the actual quality of care due to lack of technical expertise or lower expectations. Despite that, we tried to additionally examine objective measures of technical quality, that enable us provide more comprehensive and thus robust findings with substantial implications for clinical practice, policy and further research in such resource-limited settings.

## Conclusions

Overall, the majority (73.5%) of patients were satisfied with the services provided by the clinical laboratories. However, most of the dissatisfaction comes from the larger hospital settings. The main drivers of dissatisfaction were the cleanliness of latrine, adequacy of waiting area, ease of accessing service locations and place to put personal things, information provision, timely receipt of results and encounter of results lost, as well as the accuracy of the laboratory work. Therefore, policy-makers, health managers and providers should focus on improving the specific deficiencies related to both patient experience and technical process quality, that would have the highest impact towards enhancing the overall quality of care. Training, EQA and performance-based motivation systems should be considered to improve technical competence and efficiency of providers. In addition, redesigning the standard service ranges and client flows may need to be devised to optimize efficiency of care processes and systems, with special attention to larger hospital settings. The positive correlation observed with objective technical measures justifies importance of the patients’ perspective for driving organizational quality improvement efforts. Clinical laboratories are thus encouraged to conduct customers’ satisfaction assessments and use feedbacks in planning targeted interventions. Ultimately, further improvement efforts need to give more, and yet balanced, emphasis to customer focus, along with objective quality improvements, to reduce the disparities and enhance both satisfaction and overall care delivery.

## Data Availability

All data produced in the present study are contained in the manuscript. And raw data is also be available upon reasonable request to the authors

## Abbreviations

TAT: turnaround time of laboratory results
TB: tuberculosis
EQA: external quality assurance

## Authors’ Contributions

DDA & HBG conceived the study. DDA performed analysis and drafted the manuscript. All authors participated during the planning and conduction of the study including data collection and analysis. All authors have also reviewed and approved the final manuscript before submission.

## Authors’ Information

DDA, +251914064499

MMT, +251920199165

ATA, +251912026722

AEZ, +251913275076

SLH, +251913645489

HBG, +251913577917

## Acknowledgements

Amhara Public Health Institute (APHI) Dessie branch is dully acknowledged for funding and supporting this study. We also want our gratitude to all participants of this study including the health facilities, patients, data collectors and supervisors.

## References

1. WHO. WHO Guide for the Stepwise Laboratory Improvement Prcess Towards Accreditation in the African Region (SLIPTA). World Health Organization-Regional Office for Africa (WHO-AFRO).; 2015. 1–68 p.

2. Price CP, St John A. The Real Value of Laboratory Medicine. J Appl Lab Med. 2016;1(1):101–3.

3. WHO. Delivering quality health services: a global imperative for universal health coverage. World Health Organization, World Bank Group, OECD. Geneva: World Health Organization (WHO), OECD and The World Bank: World Health Organization (WHO), OECD and The World Bank; 2018. 1–100 p.

4. MoH. ETHIOPIAN NATIONAL HEALTH CARE QUALITY STRATEGY 2016-2020: Transforming the Quality of Health Care in Ethiopia. Federal Democratic Republic of Ethiopia, Ministry of Health (MoH), Addis Ababa, Ethiopia.; 2016.

5. WHO. Primary Health Care Systems (Primasys): Case study from Ethiopia. World Health Organization. Geneva: World Health Organization (WHO); 2017.

6. Girma M, Desale A, Hassen F, Sisay A, Tsegaye A. Survey-Defined and Interview-Elicited Challenges That Faced Ethiopian Government Hospital Laboratories as They Applied ISO 15189 Accreditation Standards in Resource-Constrained Settings in 2017. Am J Clin Pathol. 2018;150(4):303–9.

7. Grossbart SR, Agrawal J. Conceptualization and Definitions of Quality. In: Health Care Quality: The Clinician’s primer. 2012. p. 9–24.

8. Beattie M, Shepherd A, Howieson B. Do the Institute of Medicine’s (IOM’s) dimensions of quality capture the current meaning of quality in health care? - An integrative review. J Res Nurs. 2013;18(4):288–304.

9. Al-Abri R, Al-Balushi A. Patient satisfaction survey as a tool towards quality improvement. Vol. 29, Oman Medical Journal. 2014. p. 3–7.

10. Doyle C, Lennox L, Bell D. A systematic review of evidence on the links between patient experience and clinical safety and effectiveness. BMJ Open. 2013;3(1):1–18.

11. Derebe MM, Shiferaw MB, Ayalew MA. Low satisfaction of clients for the health service provision in West Amhara region, Ethiopia. PLoS One. 2017;12(6):1–10.

12. EPHI. EPHI, FMOH and ICF International. 2014 Ethiopia Service Provision Assessment Plus (ESPA+). 2014.

13. Desalegn DM, Abay S, Abebe A, Lulie AD, Dejene D, Mersha TB, et al. Quality of Focused Antenatal Care Laboratory Services Provided at Public Health Facilities in Addis Ababa, Ethiopia. Qual Prim Care. 2017;26(3):81–9.

14. Almasabi M, Yang H, Thomas S. A Systematic Review of the Association Between Healthcare Accreditation and Patient Satisfaction. World Appl Sci J. 2014;31(9):1618–23.

15. Tefera Z, Tsegaye A, Hassen F. Assessment of Patient satisfaction towards Clinical laboratory services among Strengthening Laboratory Management towards Accreditation (SLMTA) Program Implementing Hospital Laboratories under Addis Ababa City Administration, Ethiopia. College of Health Sciences, Addis Ababa University; 2017.

16. Barghouthi E a D, Imam A. Patient Satisfaction: Comparative Study between Joint Commission International Accredited and Non-accredited Palestinian Hospitals. Heal Sci J. 2018;12(1):1–7.

17. Donabedian A. The Definition of Quality and Approaches to Its Assessment. Health Serv Res. 1981;16(2):236.

18. World Health Organization. Toolkit on monitoring health systems strengthening - Service Delivery (Draft). World Health Organization (WHO).; 2008. p. 1-18.

19. Donabedian A. The quality of care: How can it be assessed? JAMA J Am Med Assoc. 1988;260(12):1743–8.

20. Mosadeghrad A. A Conceptual Framework for Quality of Care. Mater Socio Medica. 2012;24(4):251.

21. Chawla R, Goswami B, Singh B, Chawla A, Gupta VK, Mallika V. Evaluating laboratory performance with quality indicators. Lab Med. 2010;41(5):297–300.

22. Hailu HA, Desale A, Yalew A, Asrat H, Kebede S, Dejene D, et al. Patients’ satisfaction with clinical laboratory services in public hospitals in Ethiopia. BMC Health Serv Res. 2020;20(1):1–9.

23. Gill L, White L. A critical review of patient satisfaction. Leadersh Heal Serv. 2009;22(1):8–19.

24. Wisniewski JM, Diana ML, Yeager VA, Hotchkiss DR. Comparison of objective measures and patients’ perceptions of quality of services in government health facilities in the Democratic Republic of Congo. Int J Qual Heal Care. 2018;30(6):472–9.

25. Farley H, Enguidanos ER, Coletti CM, Honigman L, Mazzeo A, Pinson TB, et al. Patient satisfaction surveys and quality of care: An information paper. Ann Emerg Med. 2014;64(4):351–7.

26. Alhassan RK, Duku SO, Janssens W, Nketiah-Amponsah E, Spieker N, Van Ostenberg P, et al. Comparison of perceived and technical healthcare quality in primary health facilities: Implications for a sustainable National Health Insurance Scheme in Ghana. PLoS One. 2015;10(10):1–19.

27. Alelign A, Belay YA. Patient satisfaction with clinical laboratory services and associated factors among adult patients attending outpatient departments at Debre Markos referral hospital, Northwest Ethiopia. BMC Res Notes. 2019;12(1):1–6.

28. Abera RG, Abota BA, Llegese MH, Nnegesso AE. Patient satisfaction with clinical laboratory services at tikur anbessa specialized hospital, Addis Ababa, Ethiopia. Patient Prefer Adherence. 2017;11:1181–8.

29. Assefa F, Mosse A, H/Michael Y. Assessment of Clients’ Satisfaction with Health Service Deliveries at Jimma University Specialized Hospital. Ethiop J Health Sci. 2011;21(2):101–9.

30. Péfoyo AJK, Wodchis WP. Organizational performance impacting patient satisfaction in Ontario hospitals: a multilevel analysis. BMC Res Notes. 2013;6(509):1–12.

31. Price RA, Elliott MN, Zaslavsky AM, Hays RD, Lehrman WG, Rybowski L, et al. Examining the role of patient experience surveys in measuring health care quality. Med Care Res Rev. 2014;71(5):522–54.

32. Park DA. Client Satisfaction Evaluation. Vol. 8, Employee Assistance Quarterly. Geneva: World Health Organization (WHO). WHO/MSD/MSB 00.2g; 1993.

33. ANRSHB. Amhara National Regional State Health Bureau Comprehensive Laboratory Guideline. Amhara National Regional State Health Bureau (ANRSHB); Bahir Dar, Ethiopia; 2016.

34. Turner AG, Angeles G, Tsui AO, Wilkinson M, Magnani R. Sampling Manual for Facility Surveys: for Population, Maternal Health, Child Health and STD Programs in Developing Countries. MEASURE Evaluation Manual Series, No.3. MEASURE Evaluation. Carolina Population Center, University of North Carolina at Chapel Hill.: MEASURE Evaluation.; 2001. 1-146 p.

35. Tegegne B, Ejigu K, Alemu G, Fetene Y, Endaylalu K, Melese M. Performance of malaria microscopy external quality assessment and networking among health facilities in west Amhara region, Ethiopia. BMC Infect Dis. 2020;20(1):1–6.

36. Hailu HA, Shiferaw MB, Demeke L, Derebe MM, Gelaw ZD, Emiru MA, et al. External quality assessment of malaria microscopy diagnosis among public health facilities in West Amhara Region, Ethiopia. BMC Res Notes. 2017;10(1):1–5.

37. Ambelie YA. Patients’ Satisfaction and Associated Factors among Private Wing Patients at Bahirdar Felege Hiwot Referral Hospital, North West Ethiopia. Sci J Public Heal. 2014;2(5):417.

38. Abdosh B. The quality of hospital services in Eastern Ethiopia: Patients’s perspective. Ethiop J Heal Dev. 2009;20(3):199–200.

39. Mekonnen A, Mariam ZT, Kedir H. Patient Satisfaction with Laboratory Services in Selected Government Hospitals, Eastern Ethiopia. Harar Bull Heal Sci. 2011;1(3):12–24.

40. Yeshanew AG, Geremew RA, Temesgen MK. Assessments of patient and health care workers satisfaction on the laboratory services in St. Paul’s hospital millennium medical college, Addis Ababa, Ethiopia. Int J Sci Reports. 2017;3(7):192.

41. Tadele G, Ejeta E, Desalegn M, Abere S, Elias K. Patients Satisfaction on Clinical Laboratory Services at Nekemte Referral Hospital, Oromia, Ethiopia. Food Sci Qual Manag. 2014;30(16):25–31.

42. Mindaye T, Taye B. Patients satisfaction with laboratory services at antiretroviral therapy clinics in public hospitals, Addis Ababa, Ethiopia. BMC Res Notes. 2012;5.

43. Teklemariam Z, Mekonnen A, Kedir H, Kabew G. Clients and clinician satisfaction with laboratory services at selected government hospitals in eastern Ethiopia. BMC Res Notes. 2013;6(1):1–7.

44. Belay M. HIV/AIDS Patients′ Satisfaction on ART Laboratory Service in Selected Governmental Hospitals, Sidamma Zone, Southern Ethiopia. Sci J Public Heal. 2013;1(2):85.

45. Bogale AL, Kassa HB, Ali JH. Patients’ perception and satisfaction on quality of laboratory malaria diagnostic service in Amhara Regional State, North West Ethiopia. Malar J. 2015;14(1):1–7.

46. Hu R, Liao Y, Du Z, Hao Y, Liang H, Shi L. Types of health care facilities and the quality of primary care: a study of characteristics and experiences of Chinese patients in Guangdong Province, China. BMC Health Serv Res. 2016;16(335):1–11.

47. Ochan AW, Aaron K, Aliyu S, Mohiuddin M, Bamaiyi PH. Patients’ Satisfaction with Healthcare Services Received in Health Facilities in Bushenyi District of Uganda. Int J Sci Healthc Res. 2018;3(1):76–87.

48. Haj-Ali W, Karroum LB, Natafgi N, Kassak K. Exploring the relationship between accreditation and patient satisfaction-the case of selected Lebanese hospitals. Int J Heal Policy Manag. 2014;3(6):341–6.

49. Dansereau E, Masiye F, Gakidou E, Masters SH, Burstein R, Kumar S. Patient satisfaction and perceived quality of care: Evidence from a cross-sectional national exit survey of HIV and non-HIV service users in Zambia. BMJ Open. 2015;5(12):1–11.

50. Kumar Acharya S, Kumar Sharma S, Dulal B, Kumar Aryal K. Nepal Health Sector Support Program, Ministry of Health and Population. DHS Further Analysis Reports No. 112. DHS Further Analysis Reports No. 112. Rockville, Maryland, USA: ICF.; 2018.

51. Rao M, Clarke A, Sanderson C, Hammersley R. Patients’ own assessments of quality of primary care compared with objective records based measures of technical quality of care: Cross sectional study. Br Med J. 2006;333(7557):19–22.

52. Hashem T. Patient Satisfaction Evaluation on Hospitals; Comparison Study Between Accredited and Non Accredited Hospitals in Jordan. Eur Sci J. 2015;11(32):298–314.

